# Fear of exponential growth in Covid19 data of India and future sketching

**DOI:** 10.1101/2020.04.09.20058933

**Authors:** Supriya Mondal, Sabyasachi Ghosh

**Affiliations:** Aadarsh Nursing Institute, Raipur, Chhattisgarh 492015, India; Indian Institute of Technology Bhilai, GEC Campus, Sejbahar, Raipur 492015, Chhattisgarh, India

## Abstract

We have attempted to interpret existing n-cov positive data in India with respect to other countries - Italy, USA, China and South Korea. We have mainly zoomed in the exponential growth in a particular zone of time axis, which is well followed in the data profile of India and Italy but not in others. A deviation from exponential growth to Sigmoid function is analyzed in the data profile of China and South Korea. Projecting that pattern to time dependent data of total number and new cases in India, we have drawn three possible Sigmoid functions, which saturate to cases 10^4^, 10^5^, 10^6^. Ongoing data has doubtful signal of those possibilities and future hope is probably in extension of lock-down and additional imposition of interventions.

## 1 Introduction

Presently, people of entire world is in fear from Covid19 spreading which started from wuhan, China on December 2019. Within a three months time period, the spreading from China to entire world become so violent that WHO declared it as a pandemic disease on 11th March, 2020 [1]. In present draft, we are just going to describe the existing n-cov data of India with respect to other countries where we have zoomed in the exponential growth and its corresponding time axis zone. Then, its deviation from exponential growth is discussed and we have sketched different possible deviated curves, which may be or may not be expected from present lock-down schenario of our country (India).

## 2 Identification of Exponential growth

Without any artificial immunization, the virus infection can spread exponentially [2]. A recent time example is the spreading of Covid19 infection in different countries, whose real-time documentation can be seen in world-meter data [3]. This growth alarm already gave a threat on survival probability of human civilization and force them to think many interventions to fight against this virus. These interventions can make a deviation from exponential curve, which will be reflected in real and raw data of different countries.

Here we have attempted to analyze the raw data of India with comparison to other countries and tried to understand the growth pattern. Taking the data of ncov positive cases for India, Italy, USA, South Korea (SK), China from the wikipedia links [4, 5, 6, 7, 8], we have plotted in the left panel of Fig. (1), where × axis is denoting as days and y axis is denoting the number of cases. The day, from when first cases is detected, is considered as day one. The first case detected in India, Italy, USA, SK, are 30th January, 31st January, 20th January, 20th January in 2020 respectively and for China date of first detection is not very clear but wuhan CDC admitted about a cluster of cases on 31st December, 2019. Though detection dates of first case is different for different countries but they are considered as same initial point in the graph. The y axis is taken in logarithm scale to cover rapidly growing numbers of +ve cases with days. If we assume an exponential growth of +ve cases

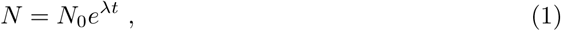

where *N*_0_ is number of case in day one, then one expect a constant value of *λ*, which can be obtained from the relation

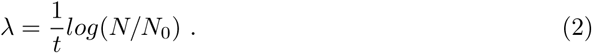

**Figure 1:**
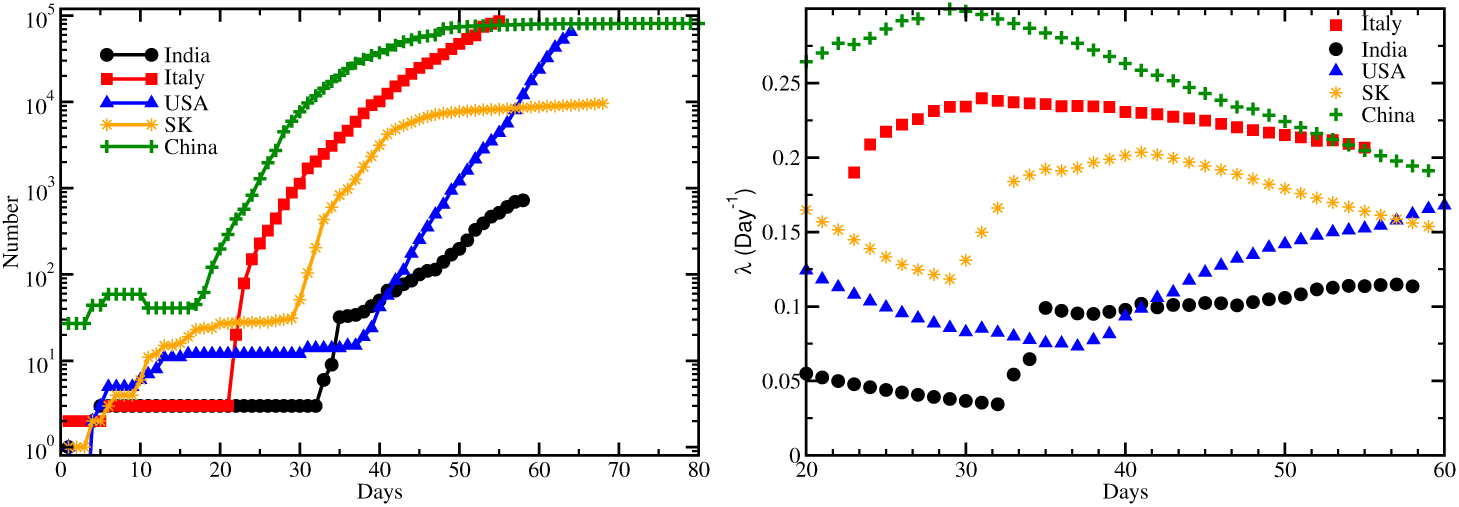
Left panel: No of + ve cases in India (black circles), Italy (red squares), USA (blue triangles), SK (brown stars), China (green pluses) with days. Right panel: Assuming exponential growth of positive ncov cases, *λ* vs days (keeping *N*_0_ = 1).

Using the *N* (*t*), given in left panel of Fig. (1), we have obtained *λ* from the Eq. (2), which is plotted in the right panel of Fig. (1). In principle *λ* should be constant if the curve follows exact exponential growth but in reality it may not follow. Therefore, we get non monotonic curve of *λ* as a function of days instead of a horizontal curve. Another important point, as a simplified model, we have considered *N*_0_ = 1 for each data points to see the approximate deviation in growth of cases.

If we critically analyze the left panel of Fig. (1), then we can identify a threshold date of different countries, after which a rapid growth is started. Here we see the day 32, 21, 37, 29, 17 may be roughly considered as the threshold day for India, Italy, USA, SK, and China respectively. At that point of time, their respective number of cases were 3, 3, 14, 33, 46. However if we are interested to identify the exponential growth then we have to focus on particular time axis zone, where a linear growth in our logarithmic figure (left panel of Fig. (1)) is observed. For India and Italy, we notice this linear growth in log scale from *t*_0_ =day 35 and 24. Right panel of Fig. (1) support this exponential growth by showing approximately constant values of *λ* in the range day 35-57 (5th - 27th March) and day 24- 56 (24th Feb - 27th March) for India and Italy respectively. On day 35 for India-data and day 24 for Italy-data the number of cases are 31 and 150, which are considered as *N*_0_ of exponential growth function, described in Eq. (1). For India, Italy, we can find the exponential relation with *λ* = 0.11, 0.22 by starting time axis from 5/March/20 and 24/Feb/20 respectively. If we noticed the right panel of Fig. (1), then we can see that after 5/March/20 and 24/Feb/20 i.e. after day 35 and 24 the *λ* remain constant (0.11 and 0.22) for India and Italy. Although, we should remember that approximate constant *λ* = 0.11, 0.22 within day 35-58, 24-56 for India, Italy are obtained when we crudely consider *N*_0_ = 1. Actually we have to do exponential fitting within day 35-58, 22-56 with initial values *N*_0_ = 31, 150 as shown in left panel of Fig. (2). For India, *N*_0_ = 31, *λ* = 0.14 can able to fit data within day 35-58. Hence, on 27th March or day 57, we will get 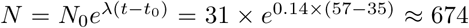, which is close to exact data 694. On the other hand, Italy-data within day 24-56 can be fitted by two sets - *N*_0_ = 150, *λ* = 0.31 in day 24-34 and *N*_0_ = 3500, *λ* = 0.15 in day 34-56. So, on 5th March or day 34, *N* = 150 × *e*^0.31×(34− 24)^ ≈3, 329, which is close to exact data 3, 089 and next, on 27th March or day 56, *N* = 3300 × *e*^0.15×(56− 34)^ ≈ 89, 471, which is close to exact data 86, 498. These information are briefly tabulated in Table (1). Extrapolating the last exponential growth of India and Italy is really a danger alert, but we should be fear on that extrapolation possibility until or unless the interventions, taken from those countries, will defeat this exponential growth. To cover Indian population ∼ 10^9^, required time scale can be calculated as

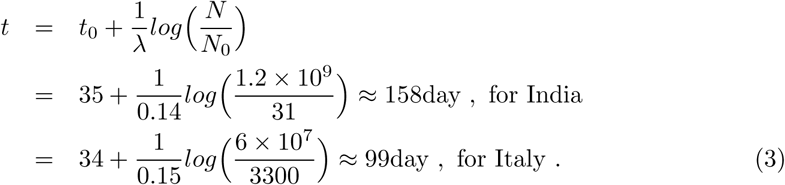

158 day means around 1st week of July for India and 99 day means around 1st week of May for Italy. It means that next each dates are vital to us and we should adopt all possible interventions, by which we can transform this exponential growth to a very stable and mild growth function like Sigmoid function [9]. Recent time Refs. [10, 11, 12] might be a guiding points about that kind of studies. Next section will focus on that kind of discussions.

**Figure 2:**
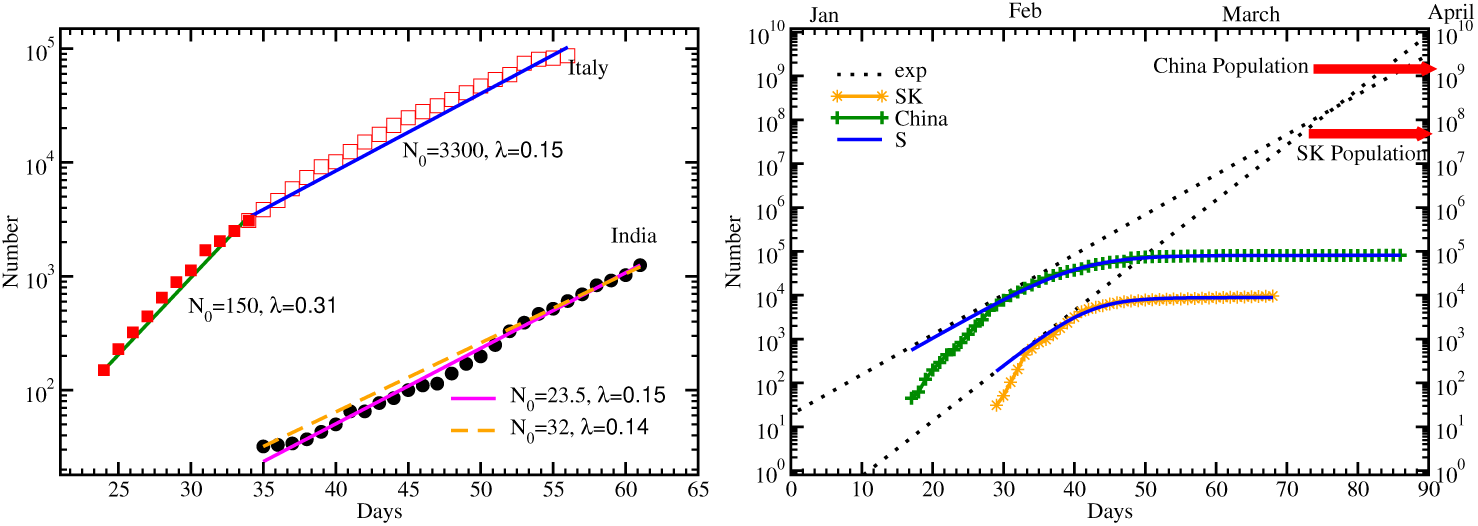
Left panel: Identifying the time zone - day 35 − 57 for India and day 24-56 for Italy, exponential fitting has been done. Right panel: Fitting the data of China (pluses) and SK (stars) by Sigmoid-type function and drawing their possible exponential growth, from where a fruitful deviation has been made due to their undertaken interventions.

**Table 1:**
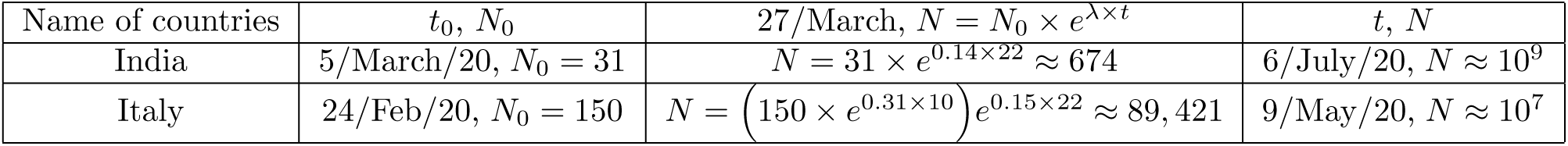
Identifying exponential growth via different time and number of cases for India and Italy.

## 3 Deviation from Exponential growth and Future sketching

The deviation of exponential function is really necessary to prevent this massive infection spread. The *λ* is main controlling parameter, which is a proportional constant for the relation 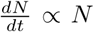, which means no. of new cases ∝ total no cases. In real data this proportional relation does not hold for entire time axis, therefore we find a time dependent *λ* Instead of a constant value. However, we can identify different time zones, within where *λ* become constant and from one zone to other this constant value changes. Positive changes is not a good news for us but negative changes is a hope for possibility of transformation this exponential function to Sigmoid-type function.

Let us assume that our real data, carry a time dependent *λ*(*t*), which may be splitted into time-independent (*λ*_0_) and dependent part (Δ*λ*(*t*)) as

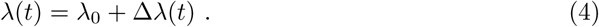

So real data (RD) follow the relation

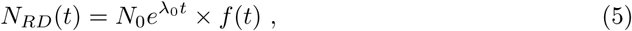

where *f* (*t*) = *e*^Δ*λt*^ is an important function, which can suppress the exponential growth for negative values of Δ*λ*(*t*). If Δ*λ* = 0, then we will get exactly exponential function

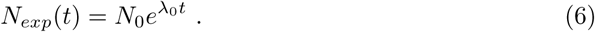

Through proper interventions, there is a possibilities of getting the time dependent functions,

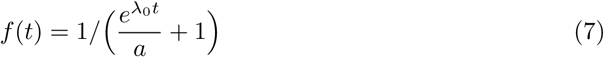

i.e.

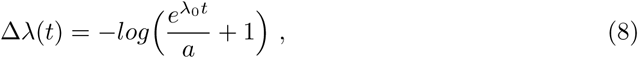

for which our exponential function will transform to a Sigmoid function

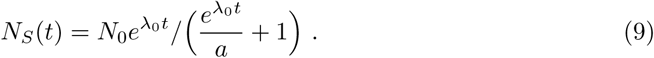

Here *a* is very important parameter which fix the maximum number of cases *N*_*max*_ = *N*_0_ × *a*, where the sigmoid curve will saturate.

As we found that China and SK data already reached to a Sigmoid-type function, so they may be used as good example where transformation of exponential curve to sigmoid functions has been achieved. In right panel of Fig. (2) we have attempted to fit the data of China and SK via sigmoid function (solid line), given in Eq. (9), and we obtained the parameters - *λ*_0_ = 0.21, *a* = 177.8 and *λ*_0_ = 0.29, *a* = 46.6 respectively. We know that for small *t*, sigmoid function cannot be distinguishable from exponential function, because *f* (*t*) → 1, when 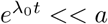, or in other word - when *t* is very small. Therefore, we draw the 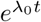 curve(dotted line) for China and SK data in the right panel of Fig. (2) and we noticed the merging of exponential and Sigmoid function in low *t* zone. Pointing out China and SK populations by red arrows, the exponential growth might reach within April-May.

The countries, which have not achieved the sigmoid type function pattern and still continuing the exponential growth, should follow proper interventions to turn into sigmoid function (*N*_*S*_) from exponential function (*N*_*exp*_). For India, projecting the total number of cases data (black triangles) we have drawn exponential curve (solid orange line) with *N*_0_ = 31, *λ*_0_ = 0.14 and different possible Sigmoid functions - *S*_1_ (blue dash line), *S*_2_ (red dotted line), *S*_3_ (green dash-dotted line) in upper left panel of Fig. (3) whose zoomed version is repeated in it’s upper right panel. The *S*_1_, *S*_2_, *S*_3_ are designed by restricting the saturate values of number of cases - *N*_*max*_ = *N*_0_ × *a* = 10^4^, 10^5^, 10^6^ respectively. Similarly, projecting the new cases data (black circles), we have drawn time derivative of different Sigmoid functions in lower left panel of Fig. (3) whose zoomed version is repeated in it’s lower right panel.

**Figure 3:**
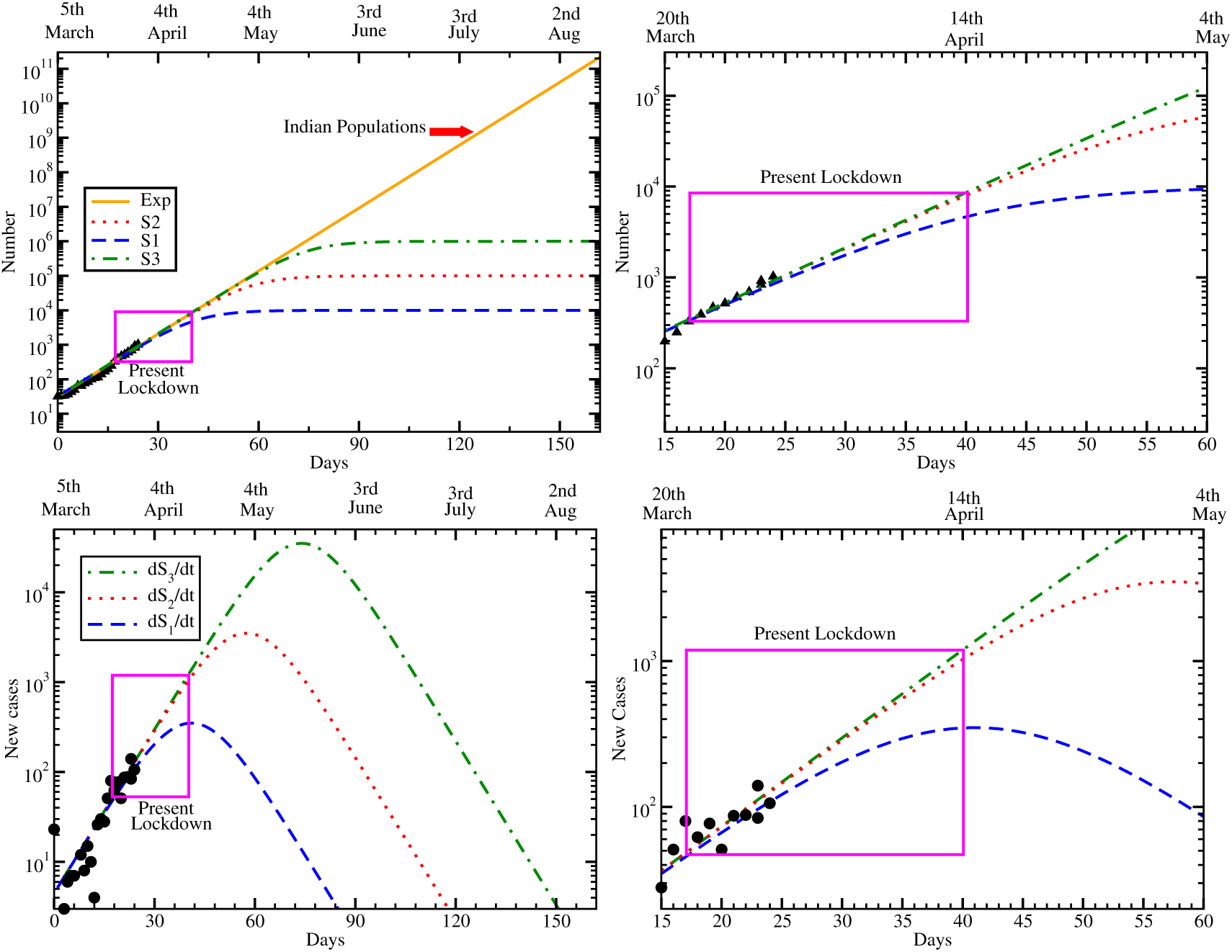
(a) Upper left panel: To match total case data of India, the exponential function (brown solid line), and different possible sigmoid functions *S*_1,2,3_ (blue dash, red dotted and green dash-dotted lines). (b) Upper right panel: Zoom version of (a) to see the association of present lock-down (denoted by box). (c) Lower left panel: To match new case data of India, time derivative of different possible sigmoid functions *S*_1,2,3_ (blue dash, red dotted and green dash-dotted lines). (d) Lower right panel: Zoom version of (c) to see the association of present lock-down (denoted by box).

In Fig. (3), data are taken from 5th March from when exponential growth is started. Within 1st week of July this exponential growth might cover the entire Indian population (10^9^), pointed by red arrow in upper left panel of Fig. (3). Till now Covid-19 containment is not possible because vaccine is not available to stop the viral spreading. So we have to emphasize on mitigation (measures taken to slow it’s spreading) measures/ interventions like lock-down; screening, testing and isolation of mass population; hand hygiene; using mask etc. Indian Govt declared lock down for 21 days from 23rd March to 14th April which is denoted by pink box in Fig. (3). In the upper right panel of Fig. (3) we can see that data is till not achieved any sigmoid functions so it may be considered as part of exponential function or low *t* limit of sigmoid function. To achieve *S*_1_ function with *N*_*max*_ = 10^4^ is quite challenging task to our country. Within the lock-down box, if we can see a rapid suppression then it is possible, otherwise we have to hope on *S*_2_ and *S*_3_ with *N*_*max*_ = 10^5^ and 10^6^, whose rapid deviation from exponential curve will be seen after lock-down period. It is only possible after extending the lock-down with more additional interventions. Hence coming data are very important to us and more critical analysis is required on those data.

Another important quantity is new cases, which is plotted in lower panel of Fig. (3). Approximately time derivative of total cases will give new cases as a function of time. For exact exponential growth, given in Eq. (6), we will get time derivative:

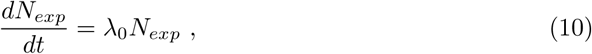

while for Sigmoid function, given in Eq. (9), we will get time derivative:

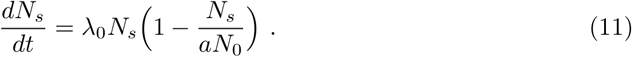

So our expectation is to transform the Eq. (10) to (11) in new cases data. The zoomed version of new cases graph, shown in right panel of Fig. (3), is supporting our earlier conclusion obtained for total cases data, discussed above.

## 4 Summary and Conclusion

In summary, we have attempted to identify growth pattern of covid19 cases in India, where other country like Italy, USA, China and South Korea are also considered for reference point of our understanding. Realistic data of different countries carry their own complexity, but still all of them has common trend that initially they follow very mild growth and then suddenly a rapid growth. After that rapid growth, they have a tendency to follow exponential pattern, which is well maintained in the data of India and Italy but the data of USA, China and South Korea follow very dynamical growth. Present article is intended to zoom in the straight forward alarm that exponential growth can cover entire population of India (Italy) within July (May) if it will be continued. In this regards, a positive hope can be found from the data profile of China and South Korea, whose transformation pattern from exponential growth function to a stable Sigmoid function is sketched and analyzed. As a positive hope from present interventions, mainly lock-down, considered by India Govt., we have sketched three possible Sigmoid functions, which can be saturated in either 10^4^ or, 10^5^ or, 10^6^. Within the lock-down period, we have not found any deviation trend from exponential to Sigmoid function till now. A rapid, intermediate and mild reduction can create a possibility of saturation within 10^4^, 10^5^ and 10^6^ but all of them never be achieved probably without extending lock-down. Apart from them, we might have to think about additional interventions for getting deviated from exponential to Sigmoid-type growth.

## Data Availability

Cited in references

https://www.covid19india.org/

## Acknowledgment

SM and SG thank to their daughter Adrika Ghosh for allowing time for this investigation during lock-down period.

